# Twin study provides heritability estimates for 2,321 plasma proteins and assesses missing SNP heritability

**DOI:** 10.1101/2024.04.24.24306270

**Authors:** Gabin Drouard, Fiona A. Hagenbeek, Miina Ollikainen, Zhili Zheng, Xiaoling Wang, FinnGen, Samuli Ripatti, Matti Pirinen, Jaakko Kaprio

**Author notes:** **Corresponding authors:** Gabin Drouard & Jaakko Kaprio. A complete list of the members of the FinnGen banner can be found in the supplementary material.

## Abstract

Assessing how much of the variability in blood plasma proteins is due to genetic or environmental factors is essential for advancing personalized medicine. While large-scale studies have established SNP-based heritability (SNP-h^2^) estimates for plasma proteins, less is known about the proportion of total genetic effects on protein variability. We applied quantitative genetic twin models to estimate the heritability of 2,321 plasma proteins and to assess the proportion of heritability accounted for by SNP-h^2^ estimates. Olink proteomics data were generated for 401 twins aged 56-70, including 196 complete same-sex twin pairs. On average, 40% of protein variability was attributable to genetic effects. Twin-based heritability estimates were highly correlated with published SNP-h^2^ estimates from the UK Biobank (Spearman coefficient: r=0.80). However, on average, only half of the total heritability was covered by SNP-h^2^, and the other half, representing one-fifth of total protein phenotypic variability, remains missing.

---

Major financial and human resources have been devoted to genotyping populations, resulting in well-known large biobanks such as the UK Biobank^2^ or FinnGen^3^. As a result, identifying new drug targets has become a great opportunity, opening the door toward personalized medicine. While the knowledge of the interplay between genes and health has increased dramatically in recent years, the understanding of how genetic information translates into health status remains to be explored. Further study of the proteome could provide a new functional perspective on disease and better link genotypes to phenotypes^4^.

Protein information has long been available and used in biomedical research, however, proteomic studies have mainly been hypothesis-driven and applied to only a handful of proteins, leaving most of the proteome undiscovered. Advances in several high-throughput technologies have enabled the identification of more proteins with high accuracy^4^; mass spectroscopy (MS) being one of them. MS has been largely democratized within academic laboratories enabling the measurement of hundreds to thousands of proteins in biosamples, typically derived from blood.

While academic platforms have been the initial drivers of this revolution, variability in the instrumentation and software used by individual academic proteomic centers has hindered the replication and comparability of results from different studies. With the aim of unifying and standardizing proteomics production, commercial companies, such as Olink and SomaLogic, have developed their own products. As a result, large cohorts and biobanks have undertaken large-scale determinations of the proteome, such as the UK Biobank which recently generated Olink proteomic data on approximately fifty thousand individuals^1^.

One feature that needs to be characterized for large-scale proteomic studies is the relative contribution of genetic and environmental effects to protein variability. Recently, large-scale proteomic studies in the UK Biobank have identified thousands of genetic associations with protein levels and common traits^1,5^. Highly heritable proteins are unlikely to be substantially influenced by environmental factors, and thus may not be efficient targets for interventions.

Conversely, proteins that show little genetic influence on their variability may be ideal for intervention, but may not be optimal for causal investigations utilizing genetic variants^6^. Polygenic risk scores^7^ (PRSs) for proteins with too little variability due to genetic influences may also be inadequate for personalized risk assessment in clinical settings. Determining the extent to which genetics and environment influence protein variability is therefore of great importance.

Recently, a study from the UK Biobank^1^ estimated SNP heritability (SNP-h^2^) for 2,923 proteins generated with the Olink platform. However, SNP-h^2^ estimates for protein levels provide only partial knowledge of how genetics influence protein variability, as common variants account for only a fraction of the total genetic influences on traits^5^. Twin cohorts could, yet, allow quantification of the total genetic effects on protein variability, should the effects of genes be additive or non-additive. Twin-based heritability estimates can be obtained using classical twin modeling, which relies on the comparison between monozygotic (MZ) and dizygotic (DZ) twins, the former sharing 100% of their genetics with their siblings, whereas DZ twins share on average only half^8^. As such, classical twin models can assess additive genetic effects (A), dominant genetic (i.e., non-additive) effects (D), and shared (C) and non-shared (i.e., unique) environmental effects (E) on trait variability (V). One twin study^9^ has been conducted to quantify the heritability of blood proteins, but the sample size (36 MZ and 22 DZ twin pairs) and the number of proteins examined (N=342) were limited. Other twin studies have quantified protein heritability only for proteins associated with specific traits^10,11^, but, to our knowledge, no twin study has quantified protein heritability across a large number of proteins in the proteome in moderate-to-large samples.

Our study aims to fill the aforementioned gap in the literature by quantifying the total genetic influence on protein variability of 2,321 plasma proteins using a twin cohort (Figure 1). Plasma proteomic data were generated on the Olink platform for 401 twins whose ages ranged from 56 to 70 years, including 117 MZ and 80 DZ twin pairs. Twin-based heritability estimates were calculated using univariate classical twin models. We then compared these estimates with published SNP-h^2^ estimates from the UK Biobank^1^. This allowed us to estimate the proportion of the total genetic effect on protein variability accounted for by SNP-h^2^ estimates, and thus evaluate missing heritability. In addition, we also compared our twin-based heritability estimates with published PRSs of protein levels^12^ to assess whether the strength of these PRSs is increased in highly heritable proteins.

**Figure 1:**
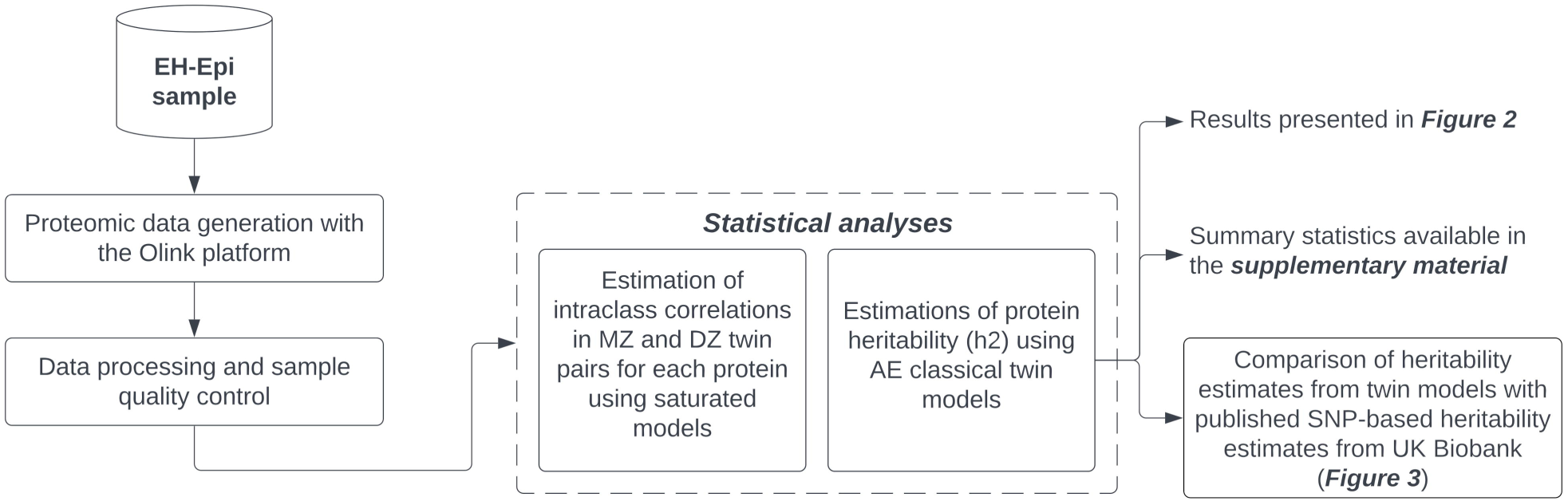
Study summary. **Caption:** Proteomic data was generated for a complete set of 401 twins. Plasma protein heritability was quantified using classical twin models for 2,321 proteins and these estimates were later compared with published SNP-based heritability estimates derived from the UK Biobank [1]. MZ: Monozygotic. DZ: Dizygotic.

We used univariate classical twin models to assess genetic effects (A) on protein variability (V) for 2,321 plasma proteins, from which heritability estimates were calculated (h^2^=A/V).

Heritability estimates varied widely (Figure 2A), with an interquartile range of 20.1% to 58.6%. Forty proteins had heritability estimates greater than 90%. The mean heritability across all plasma proteins was 40.4% with a standard deviation of 23.5%. Consequently, environmental effects accounted for 59.6% of protein variability on average. Heritability estimates for plasma proteins in the *Inflammation* and *Cardiometabolic* panels had larger median heritability estimates than those in the *Neurology* and *Oncology* panels (Figure 2B). Heritability estimates with 95% confidence intervals as well as MZ and DZ twin correlations are available in the supplementary material (Supplementary Table 1).

**Figure 2:**
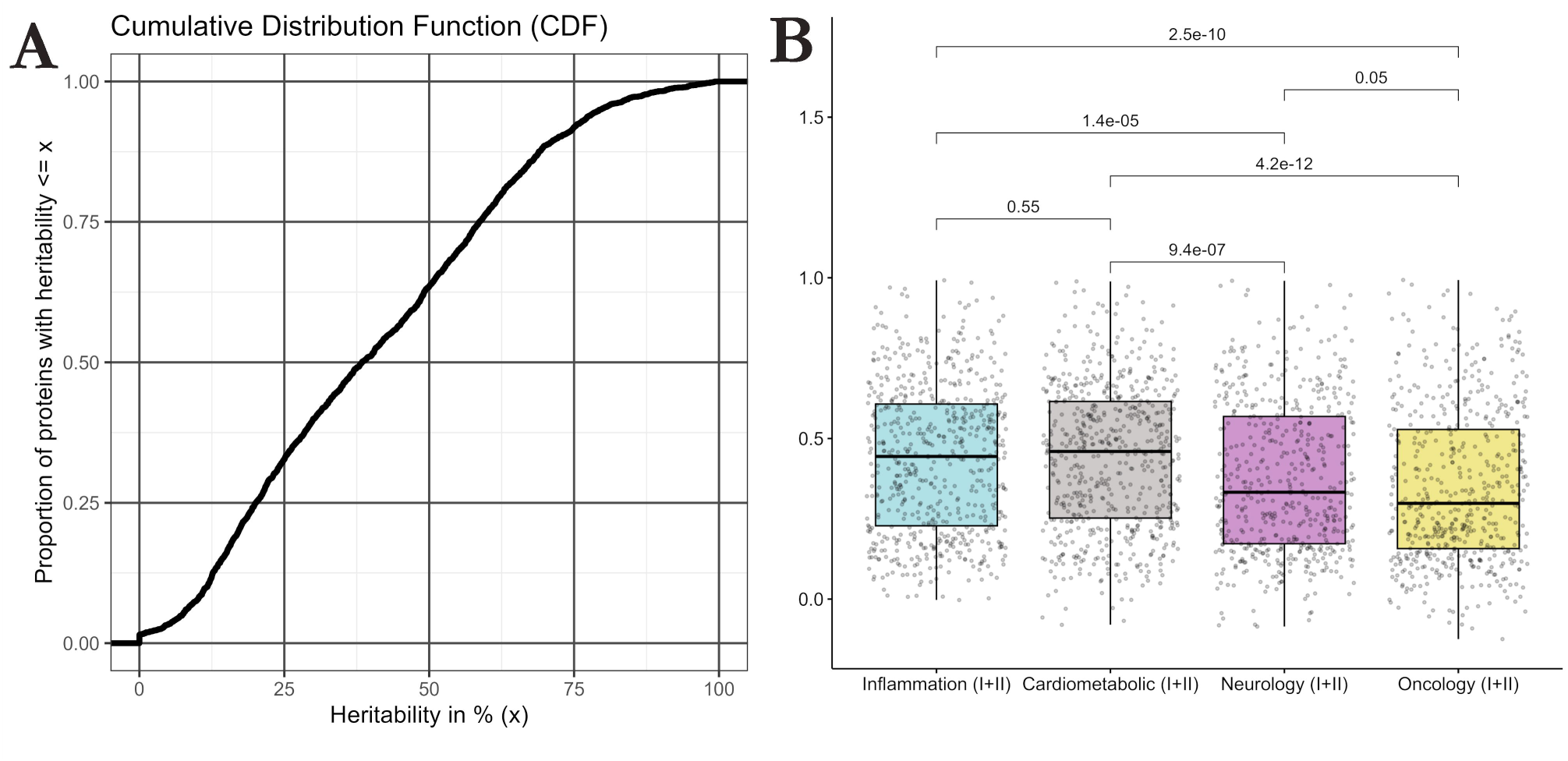
Cumulative distribution of heritability estimates derived from classical twin modeling across the entire measured proteome (A) and main Olink panels (B) **Caption:** (A) Cumulative Distribution Function (CDF) showing the proportion of proteins with heritability estimates below a value *x*, with *x* ∊ [0% , 100%]. The mean heritability of the 2321 plasma proteins was 50.4%. (B) Box plots showing the distribution of heritability estimates grouped by Olink panels. Wilcoxon tests indicated that the *Inflammation* and *Cardiometabolic* panels had proteins with greater median heritability than the *Neurology* and *Oncology* panels.

We compared the heritability estimates derived from the twin models with published SNP-based heritability estimates established in approximately 50,000 UK Biobank participants^1^. Of the 2,321 plasma proteins for which we provided heritability estimates, 2,182 overlapped with those of Sun et al.^1^. The heritability estimates for these overlapping proteins were highly correlated (Spearman correlation coefficient: ⍴= 0.80) (Figure 3A), suggesting high concordance between SNP-based and twin-based heritability estimates. However, on average, twin-based heritability estimates were higher than SNP-based estimates, as twin models estimated protein heritability to be on average twice higher (mean h^2^=40.4%) than their SNP-based counterparts (mean h^2^=20.9%) (Figure 3B). Sun et al. stated that cis primary pQTLs accounted, on average, for about one-fifth of SNP-based heritability^1^. Our study therefore suggests that identified primary pQTLs from the UK Biobank could account, on average, for approximately 10% of the total heritability quantified by twin models. In addition, the association between SNP-based and twin-based heritability estimates showed a significant non-linear fit (standardized quadratic coefficient: -15.8; p=1.4e-54), and the difference between twin-based and SNP-based heritability estimates was decreased for highly heritable proteins (Figure 3A).

**Figure 3:**
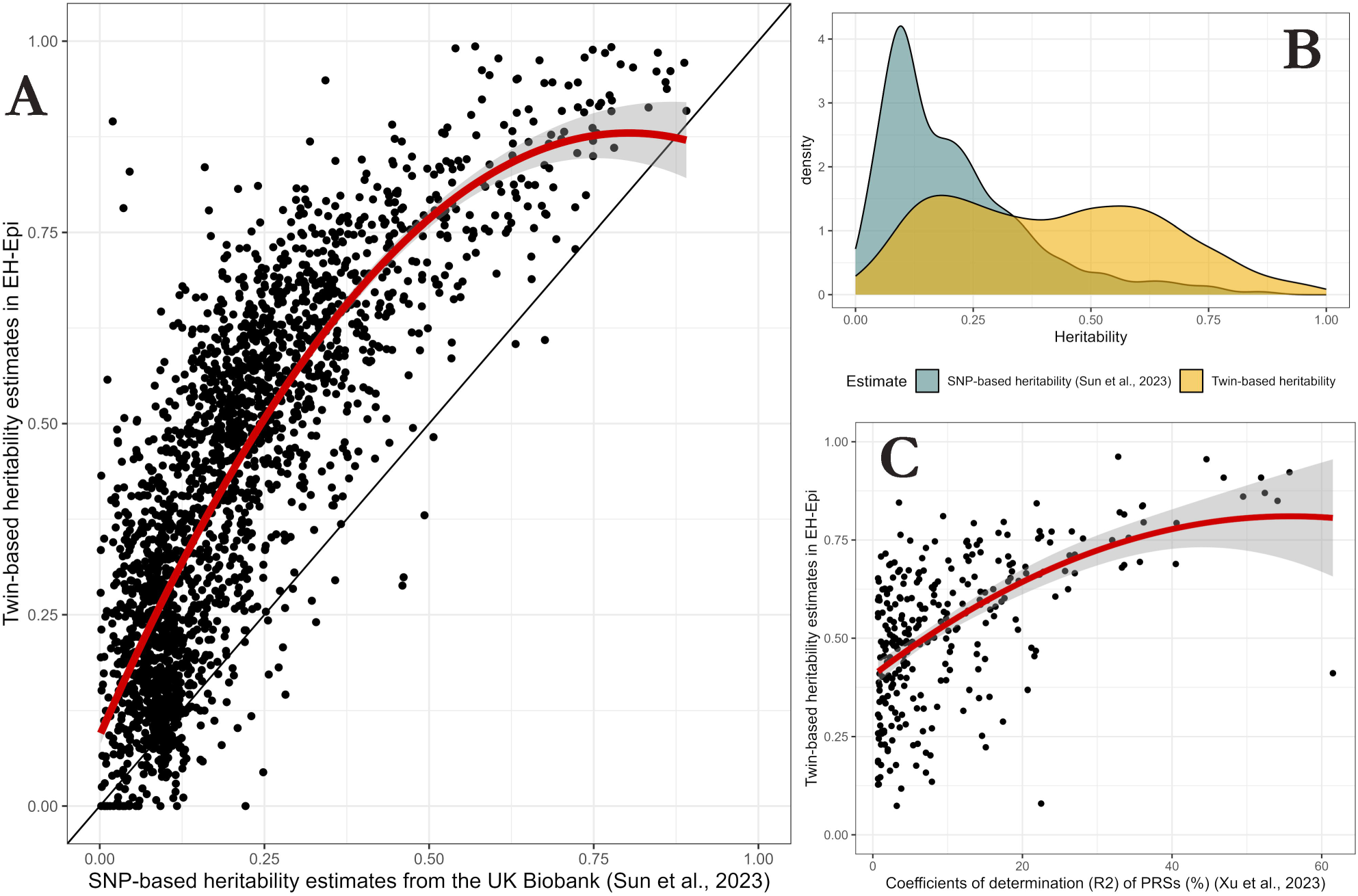
Comparison of twin-based heritability estimates with published summary statistics. **Caption: (A)** Scatterplot comparing twin-based heritability estimates with published SNP-based heritability estimates [1] in 2,182 overlapping proteins. The Spearman correlation coefficient ⍴ was equal to 0.80, indicating high agreement between heritability estimates. Proteins, represented by points, that are above the solid line “*y=x”* are proteins with higher heritability estimates from twin models than their SNP-based counterparts. A quadratic linear model was fitted (shown in red) to test for quadratic effects in the relationship between twin-based and SNP-based heritability estimates. The significant quadratic term indicates that the more heritable a protein, the closer the twin-based and SNP-based heritability estimates are. **(B)** Distribution of twin-based and SNP-based heritability estimates. Heritability estimates from twin models are more dispersed in the [0,1] segment and have higher means. **(C)** Scatterplot comparing twin-based heritability estimates with the coefficient of determination (R^2^) of the genetic scores of 294 overlapping proteins from Xu et al. [10]. A quadratic linear model was fitted (shown in red) to test for quadratic effects.

Finally, we assessed whether the genetic scores of the Olink plasma proteins published in Xu et al.^12^ correlated more strongly with proteins with high heritability estimates. For this, we used published R-squared (R^2^) coefficients of determination when the PRSs were used to predict the protein levels^12^. We quantified the correlation between twin-based heritability estimates and the R^2^ values for 294 overlapping proteins out of 308 proteins with PRS scores. The Spearman correlation coefficient was ⍴=0.54, indicating that PRSs better capture protein variability for highly heritable proteins than for low heritable proteins (Figure 3C).

In summary, our study provides heritability estimates for 2,321 blood plasma proteins derived from classical twin models. Heritability estimates averaged 40%, with about half of the proteins’ heritability estimates ranging from 20% to 59%. These estimates were on average twice as large as published SNP-based heritability estimates from the UK Biobank^1^. This indicates that, in a context in which shared environmental and non-additive effects are assumed to have no effect on protein variability, only half of the total genetic influences on protein variability can be explained by additive genetic effects of previously identified genetic variants. Thus, our study suggests that the current knowledge of the genetic basis of protein variability based on measured genetic variants is likely to be only halfway to a definitive, complete genetic understanding of protein variability.

The main limitation of our study is the use of AE twin models, from which heritability estimates are calculated while non-additive genetic (i.e., dominance) and shared environmental effects are set to zero. 1,132 out of 2,321 proteins had intra-class correlations in MZ twin pairs that were more than twice as large as those in DZ twin pairs, which suggests potential non-additive genetic effects on protein levels. Conversely, the remaining proteins (1,189 out of 2,321) could be influenced by shared environmental effects. As a result, twin-based heritability estimates of the proteins may have been inflated. Models that additionally estimate dominant genetic effects (i.e., ADE models) would allow us to separate additive from non-additive effects on protein levels, and thus assess how much heritability is missing due to additive genetic effects. Because our sample size was too modest (N=401) to assess dominant genetic effects, the missing heritability we estimated is likely due to not only additive but also non-additive genetic effects. Other limitations also need to be considered. First, the SNPs used to evaluate protein heritability were derived from participants in the United Kingdom^1^, while participants in our study are from Finland, though both populations are of European ancestry. As both SNP-based and total heritability of blood proteins may vary across ancestries, this could result in a potential over- or underestimation of missing heritability. However, confounding of age on the associations between SNPs and protein levels is relatively unlikely to affect our results, as our participants had an average age of 62, while UKB participants had an average age of 57. Larger twin studies are therefore needed to detail the genetic effects on protein variability, but also to evaluate differences in protein heritability across ancestries and age categories.

PRSs from Xu et al.^12^ were more predictive of protein levels the higher the heritability of the protein. While this could be explained by the fact that the more heritable the protein, the more genetic signal can be identified and thus the greater PRS’ R^2^, our results indicate that the gap between twin-based heritability estimates and PRS’ R^2^ was even larger for less heritable proteins, as assessed by testing for non-linearity in this association.

In conclusion, our study provides heritability estimates of plasma proteins whose magnitude indicates that genetic knowledge of protein variability in the literature is incomplete. Further efforts to separately assess the contribution of additive and non-additive genetic effects to the variability of plasma proteins are needed, both in large biobanks and in twin cohorts.

## Methods

### Cohort description

The Old Finnish Twin Cohort was initiated in 1974, and extensive questionnaire data have been collected over time^13^. Based on the wave 4 (in 2011/2012) questionnaire, twin pairs with self-reported blood pressure differing between the co-twins were identified and invited to participate in a more detailed blood pressure study on Essential Hypertension Epigenetics

(EH-Epi)^14^. The twin pairs came in for a one-day research visit where their blood pressure, height, weight, and waist circumference were measured^15^. The twins also completed interviews and questionnaires and provided fasting blood samples, from which multiple omics were generated^16,17^. In total, this EH-Epi sample included 445 twins, of whom 415 had usable plasma samples.

Proteomic data were generated using the Olink platform and after data processing (see below) a final sample of 401 twins, including 116 MZ and 80 DZ complete twin pairs, was available. Age at blood sampling and sex were included in relevant analyses along with BMI, as detailed later.

### Data processing

#### Olink proteomics

Plasma proteome profiling has been performed in the FinnGen project in several batches^3^.

One of these batches included twins from the EH-Epi sample, which we used for subsequent analyses in the present study.

Proteomic profiling was performed on 415 plasma samples of 120 mL each. The samples were analyzed using an antibody-based technology (Olink Proteomics AB, Uppsala, Sweden).

Proteomics was performed using the Olink Explore 384 (version 3.0.5) cardiometabolic (I/II), inflammation (I/II), neurology (I/II) and oncology (I/II) panels. Several protein assays did not meet the quality control criteria for Olink batch release and were excluded. These were: *BMP6*, *EPHX2*, *PGLYRP1*, *EDEM2*, *CALY*, *ARL13B*, *ARNTL*, *BCL2L11*, *BID*, *MGLL*, *EP300*, *FGF3*, *FUOM*, *KNG1*, *ADIPOQ*, *CDHR1*, *CLSTN1*, *PSG1*, *CGA*, *EFNA1*, *HTR1B*, *KCNH2*, *STXBP1*, *YAP1*, *FLI1*, *MPI*, *EBI3_IL27*, *ANGPTL7*, *CPLX2*, *TAGLN3*, *GABARAPL1*, *NFKB2*, *CTAG1A_CTAG1B*, *OGT*, *MTHFSD*, *IFIT1*, *TNPO1*, *MAGEA3*, *SH3GL3* and *RAPGEF2*.

Data was initially presented as NPX (Normalised Protein eXpression) values in long format, where NPX is Olink’s unit for quantifying relative protein concentrations on a log_2_ scale. Control samples were excluded, and NPX values were reported as missing for rows that did not meet the Olink’s internal quality control (QC) criteria. Proteins that were detected in less than 80% of the samples were excluded. NPX values below the limit of detection (LoD) were replaced by the LoD value. Outlier samples were identified using three approaches^1^ applied separately to each panel of proteins: (1) Principal Component Analysis (PCA), (2) examination of the median, and (3) the interquartile range (IQR) of the NPX across proteins. Samples with (1) at least one of the first two standardized principal component absolute values greater than 5 standard deviations (SD) from the mean, (2) a median NPX greater or lower than 5 SD from the mean sample median, or (3) an IQR of NPX greater or lower than 5 SD from the mean IQR, were excluded. For each panel, samples that passed QC were assessed graphically to show a similar distribution of NPX values across plates. Proteomic data were transformed into a wide format, resulting in 2,321 proteins. For each protein initially identified to fail QC criteria (N = 17), the imputation of the missing values was performed using its minimum LoD (average missing value rate for these proteins: 0.5%). Thus, proteomic data were available for 401 twins, including 196 complete same-sex pairs.

### Statistical analyses

#### Classical twin modeling

We used univariate classical twin (UCT) models to assess genetic and environmental influences on protein variability. These models hinge on the fact that MZ twins in a pair are more genetically similar than DZ twins in a pair, because MZ twins share all of their genetic polymorphisms at the sequence level with their co-twins, whereas DZ twins share on average only half of their segregating genes by descent from their common parents^8,18^. Therefore, greater similarity in protein levels within MZ twin pairs compared to DZ twin pairs may indicate genetic effects in the variability of that protein. UCT models traditionally decompose a trait’s total variance (V) into variance components, including additive genetic effects (A), dominance genetic effects (D), and shared (C) and non-shared (i.e., unique) environmental effects (E). As the focus of our study was to estimate the heritability of plasma proteins and our sample size is modest, we sought to quantify only the A and E components using AE twin models, which assume C and D components to be zero^19^. Heritability (h^2^) estimates were estimated and defined as A/V, which is the proportion of genetic effects (A) contributing to the total variance (V) of a protein^19^. The proportion of unique environmental effects on protein variability was termed e^2^ and defined as E/V (i.e., e^2^=1-h^2^). Sex and age at blood sampling were added as covariates in the models, and 95% confidence intervals of standardized variance components were calculated. In addition, we ran saturated models to quantify twin correlations within MZ and DZ pairs separately using maximum likelihood. Sex and age at blood sampling were added as covariates in these analyses as well. Modeling was performed using the OpenMx package (version 2.20.7), with R version 4.1.3. When fitting the saturated and AE twin models, we allowed the variance components and twin correlations to be negative. However, for graphical representations of the results, we remapped these estimates to [0,1] by considering negative estimates to be zero.

#### Association and null testing

We used non-parametric Wilcoxon rank-sum tests to assess whether the distribution of heritability estimates differed between Olink panels. Median differences were considered significant if pairwise test p-values were below 0.05.

In addition, we calculated Spearman correlation coefficients (⍴) between the protein heritability estimates derived from twin models and the SNP-based heritability estimates published in Sun et al.^1^. Spearman correlation testing was performed to assess whether the protein PRSs published in Xu et al.^12^ better predicted the levels of those proteins that were highly heritable compared with those that showed low heritability.

Finally, we tested for nonlinearity in the association between twin-based heritability estimates and SNP-based heritability estimates from Sun et al.^1^, by fitting a regression model between heritability estimates that included a quadratic term besides a linear term. We tested the need for this quadratic term by testing its nullity, at the significance level of 0.05.

## Supporting information

FinnGen banner

## Data availability

The Finnish Twin Cohort data used in the analysis is deposited in the Biobank of the Finnish Institute for Health and Welfare (https://thl.fi/en/web/thl-biobank/forresearchers). It is available to researchers after written application and following the relevant Finnish legislation.

## Acknowledgements

Data collection of the EH-Epi sample has been supported by the Academy of Finland (Grants 265240, 263278, 308248, 312073, 336832 to JK and 297908 to MO) and the Sigrid Juselius Foundation (to MO).

We want to acknowledge the participants and investigators of FinnGen study. The FinnGen project is funded by two grants from Business Finland (HUS 4685/31/2016 and UH 4386/31/2016) and the following industry partners: AbbVie Inc., AstraZeneca UK Ltd, Biogen MA Inc., Bristol Myers Squibb (and Celgene Corporation & Celgene International II Sàrl), Genentech Inc., Merck Sharp & Dohme LCC, Pfizer Inc., GlaxoSmithKline Intellectual Property Development Ltd., Sanofi US Services Inc., Maze Therapeutics Inc., Janssen Biotech Inc, Novartis AG, and Boehringer Ingelheim International GmbH. Following biobanks are acknowledged for delivering biobank samples to FinnGen: Auria Biobank (www.auria.fi/biopankki), THL Biobank (www.thl.fi/biobank), Helsinki Biobank (www.helsinginbiopankki.fi), Biobank Borealis of Northern Finland (https://www.ppshp.fi/Tutkimus-ja-opetus/Biopankki/Pages/Biobank-Borealis-briefly-in-English. aspx), Finnish Clinical Biobank Tampere (www.tays.fi/en-US/Research_and_development/Finnish_Clinical_Biobank_Tampere), Biobank of Eastern Finland (www.ita-suomenbiopankki.fi/en), Central Finland Biobank (www.ksshp.fi/fi-FI/Potilaalle/Biopankki), Finnish Red Cross Blood Service Biobank (www.veripalvelu.fi/verenluovutus/biopankkitoiminta), Terveystalo Biobank (www.terveystalo.com/fi/Yritystietoa/Terveystalo-Biopankki/Biopankki/) and Arctic Biobank (https://www.oulu.fi/en/university/faculties-and-units/faculty-medicine/northern-finland-birth-c ohorts-and-arctic-biobank). All Finnish Biobanks are members of BBMRI.fi infrastructure (www.bbmri.fi). Finnish Biobank Cooperative -FINBB (https://finbb.fi/) is the coordinator of BBMRI-ERIC operations in Finland. The Finnish biobank data can be accessed through the Fingenious® services (https://site.fingenious.fi/en/) managed by FINBB.

## Author contributions

GD, FAH, SR, MP, and JK designed the study. MO, XW, and JK were involved in data collection. GD processed the proteomic data and performed twin analyses. GD, FAH, and JK drafted the original manuscript. All authors participated in the interpretation of the results, reviewed and revised the original draft of the manuscript, and approved the final version.

---

Table S1: Heritability estimates and influence of environmental effects on protein variability derived from classical twin models with twin correlations in monozygotic and dizygotic twin pairs.

